# Prognostic value of flow-mediated dilation and computed tomography angiography findings for determining cardiovascular risk in patients with subclinical atherosclerosis

**DOI:** 10.1101/2023.04.24.23289070

**Authors:** Hideki Kawai, Masayoshi Sarai, Kunihiko Sugimoto, Sadako Motoyama, Yoshihiro Sato, Yasuomi Nagahara, Keiichi Miyajima, Takahiro Matsuyama, Hiroshi Takahashi, Akira Yamada, Hiroyuki Naruse, Hiroshi Toyama, Yukio Ozaki, Hideo Izawa

## Abstract

**Background:** Flow-mediated dilation (FMD) tests endothelial function, and computed tomography angiography (CTA) estimates the extent of coronary atherosclerosis and plaque vulnerability. This study aimed to examine the prognostic value of combining FMD and CTA in patients with no history of atherosclerotic disease.

**Methods:** The study retrospectively examined patients who underwent CTA and FMD within 3 months between 2012 and 2020. Patients with a history of cardio-cerebrovascular disease or significant stenoses on CTA were excluded. The study endpoint was defined as major cardiac and cerebrovascular events (MACCE): a composite of cardio-cerebrovascular death, acute coronary syndrome, fatal arrhythmia, ischaemic and haemorrhagic stroke, and late revascularisation of the coronary or carotid arteries 6 months after CTA. Finally, the patients were stratified into four groups based on the following factors: FMD <6.0%, per cent atheroma volume (PAV) ≥21.0%, and the presence of high-risk plaques (HRPs).

**Results:** During a mean follow-up of 4.7 years, MACCE occurred in 19 of 154 patients (mean age: 61.0±12.9 years, 89 males). FMD, PAV, and HRPs were independent predictors of MACCE after adjusting for age, sex, and hypertension. Compared with the patients with 0 points, hazard ratios of those with 1, 2, and 3 points were 2.76 (P=0.322), 9.89 (P=0.004), and 28.43 (P<0.001), respectively. Adding FMD, PAV, and HRPs to the baseline model, including age, sex, and hypertension, improved the C-index (0.712–0.831, P=0.023).

**Conclusions:** Although FMD and CTA findings are useful for predicting cardiovascular events, their combination synergises their prognostic abilities.

## Introduction

Endothelial dysfunction occurs in people with atherosclerotic risk factors before anatomical evidence of plaque formation in the arteries is observed^1^. Flow-mediated dilation (FMD), an endothelium-dependent, largely nitric oxide-mediated dilatation of conduit arteries in response to an imposed increase in blood flow and shear stress, has been widely used as a non-invasive parameter since 1992^1^. Several studies have demonstrated the prognostic value of FMD for cardiovascular events. Meta-analyses indicate a significantly lower risk of cardiovascular events by 8–13% per percentage point increase in brachial artery FMD^2-4^.

Coronary computed tomography angiography (CTA) enables the detection of subclinical coronary atherosclerosis and quantitative analysis of plaque burden in the coronary tree^5-8^. Furthermore, coronary arterial plaques with ≤30 Hounsfield unit (HU)-densities observed on CTA correlate closely with intravascular ultrasound (IVUS)-verified necrotic cores in coronary atherosclerotic plaques. We have previously reported the prognostic value of high-risk plaques (HRPs), including low-attenuation and/or positive remodelling on CTA^9-11^.

The authors of the present study hypothesised that FMD is associated with computed tomography (CT) findings, including the coronary artery calcium score (CACS), total plaque volume of coronary arterial trees, and the presence of HRPs in patients with subclinical coronary atherosclerosis. Furthermore, combining FMD and CT findings may improve their prognostic value for long-term cardio-cerebrovascular events.

## Methods

### Patients

This study retrospectively examined patients (n=158) who underwent CTA and FMD within 3 months between January 2012 and March 2020 (Figure 1). All patients with no history of cardio-cerebrovascular diseases underwent CTA for suspected angina pectoris; hence, significant stenoses (≥70% luminal stenoses on CTA in the major vessels) were excluded. Two of the 158 patients had poor image quality, and two were lost to follow-up. The following data were collected: (1) cardiovascular risk factors (hypertension, diabetes mellitus, hypercholesterolaemia, chronic kidney disease, and smoking history) and (2) medication status. The study protocol was approved by the Institutional Review Board and Ethics Committee of Fujita Health University. The study protocol conformed to the ethical guidelines of the 1975 Declaration of Helsinki as reflected in an a priori approval by the institution’s human research committee. An opt-out method from our department’s website was applied to obtain informed consent for this study.

**Figure 1.**
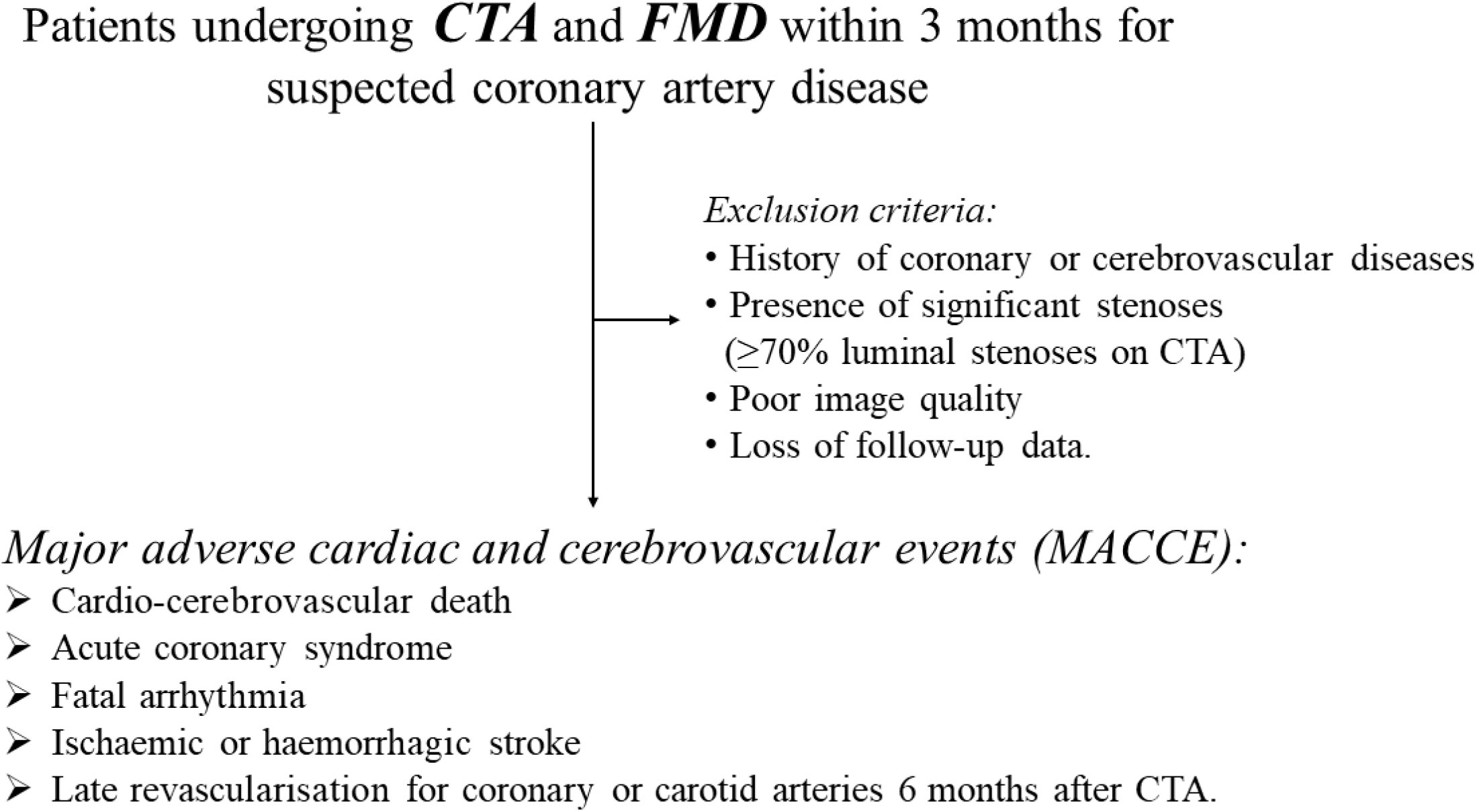
Flow chart of the study. CTA, computed tomography angiography; FMD, flow-mediated dilation

### CTA Protocol

A 320-slice CT (Aquilion ONE, Canon Medical Systems, Otawara, Japan) was performed with 0.5 mm-detector elements, rotation speeds of 350, 375, and 400 ms, and scanner settings of 350–450 mA and 100–135 kV. In patients without contraindications, oral metoprolol and/or intravenous landiolol (Ono Pharmaceutical Co., Ltd., Osaka, Japan) was administered before imaging if the heart rate was >65 beats/min. When possible, 0.4 mg sublingual nitroglycerin was administered 3–5 min before image acquisition.

For the contrast-enhanced scan, 20.4 mgI/kg/s of contrast medium was injected for 12 s, followed by 20 mL saline at 3.0 mL/s. An axial scan was performed with a prospectively gated scan in one heartbeat for heart rates <65 beats/min for half reconstruction and in two or three heartbeats for heart rates ≥65 beats/min for segment reconstruction. Raw data were reconstructed using algorithms optimised for electrocardiographic-gated reconstruction and transferred to a computer workstation for post-processing (Ziostation2, Ziosoft Inc., Tokyo, Japan)^6, 9-12^.

### CTA Analysis (Visual Assessment and Vessel Analysis)

All images were assessed by three experienced cardiologists or one radiologist (S.M., Y.N., K.M., and T.M.) blinded to each other and patients’ clinical data. The diagnosis was made by consensus in case of disagreement. Coronary artery segments with diameters >2 mm were evaluated for the presence of plaques. Coronary atherosclerotic lesions were quantified for lumen diameter stenosis by visual estimation and graded as none (0%), minimal (1–24%), mild (25–49%), moderate (50–69%), or severe (70–99%) stenoses, or occluded, according to the Society of Cardiovascular Computed Tomography guidelines^13^. The remodelling index was defined as the maximal lesion vessel diameter divided by the proximal reference vessel diameter, with positive remodelling defined as a remodelling index ≥1.1. A low-attenuation plaque was defined as any voxel with <30 HU within a coronary plaque. An HRP was defined as a plaque with positive remodelling and/or low attenuation, which are high-risk features of acute coronary syndrome (ACS), as previously reported^6, 9-12^.

Quantitative coronary atherosclerotic plaque analysis was performed by two experienced cardiologists (H.K. and Y.S.) blinded to each other, using the clinical data and visual assessment of the CT angiogram. A semi-automated plaque analysis software (QAngio CT Research Edition v2.02, Medis Medical Imaging Systems, Leiden, Netherlands) with previously validated accuracy was used. All coronary arteries with diameters ≥2 mm were evaluated using the 17-segment American Heart Association model for coronary segment classification^13^. Quantitative analysis was performed at the vessel segment level. Measurements of all segments of each vessel, including the side branches, were summed to generate the vessel length (mm), vessel volume (mm^3^), lumen volume (mm^3^), and plaque volume (mm^3^). The per cent atheroma volume (PAV) was calculated as follows: [(plaque volume/vessel volume)×100]%^6-8^.

### Brachial Artery FMD

Endothelial function was assessed non-invasively using FMD, which is the change in the brachial artery diameter after regional ischaemia. FMD was measured by an experienced sonographer (K.S.) blinded to the CT findings and clinical data. Measurements were performed according to recent guidelines by ultrasound (iE33, Philips, Amsterdam, the Netherlands) using an L11-3 Linear Probe^14-16^. Patients were instructed to fast and withhold all vasoactive medications for ≥12 h before each measurement.

The change in the brachial artery diameter was measured after 60 s of reactive hyperaemia and compared with baseline measurements after deflation to 50 mmHg higher than systolic blood pressure for 5 min, using ultrasound unit electronic callipers. Imaging was performed in a quiet, dark room. FMD was measured as the magnitude of the percentage change in brachial artery diameter from baseline to peak, and FMD rate was the maximal slope of dilation. Both measurements were performed at the same site on the right brachial artery.

### Study Endpoint and Follow-Up

Follow-up information was obtained from hospital chart reviews and supplemented with information obtained via mail (Figure 2). However, two patients were not followed up by our institution and did not reply to the mail. The study endpoint was defined as major cardiac and cerebrovascular events (MACCE), including cardio-cerebrovascular death, ACS, fatal arrhythmia, ischaemic and haemorrhagic stroke, and late revascularisation of the coronary or carotid arteries after 6 months of CTA. ACS was defined according to the fourth universal definition of myocardial infarction^17^ and the Canadian Cardiovascular Society grading of angina pectoris^18^. Strokes were defined as focal loss of neurological function caused by ischaemic or haemorrhagic events and were diagnosed by a neurologist. None of the patients underwent early coronary revascularisation 3 months after CTA.

**Figure 2.**
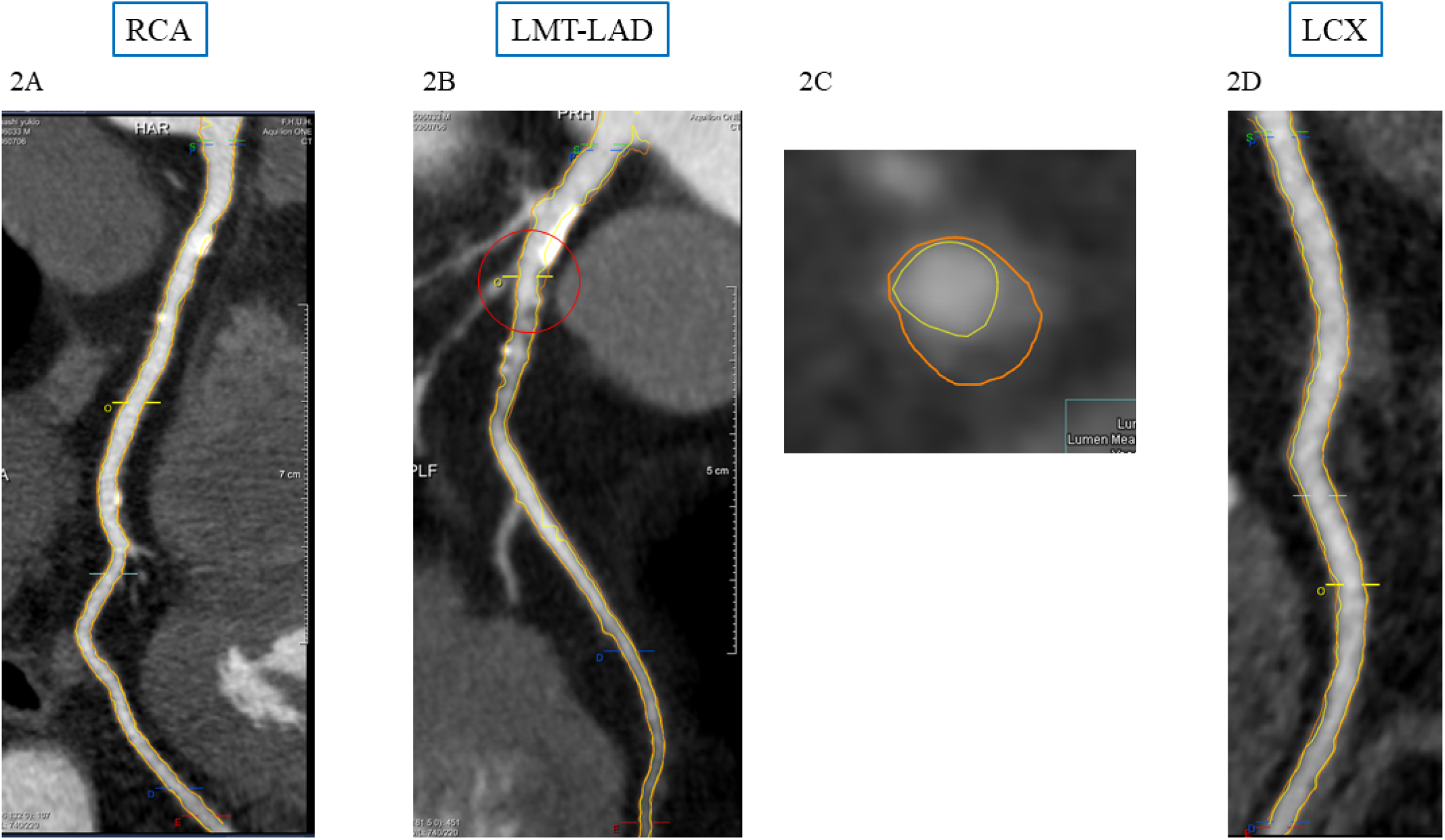
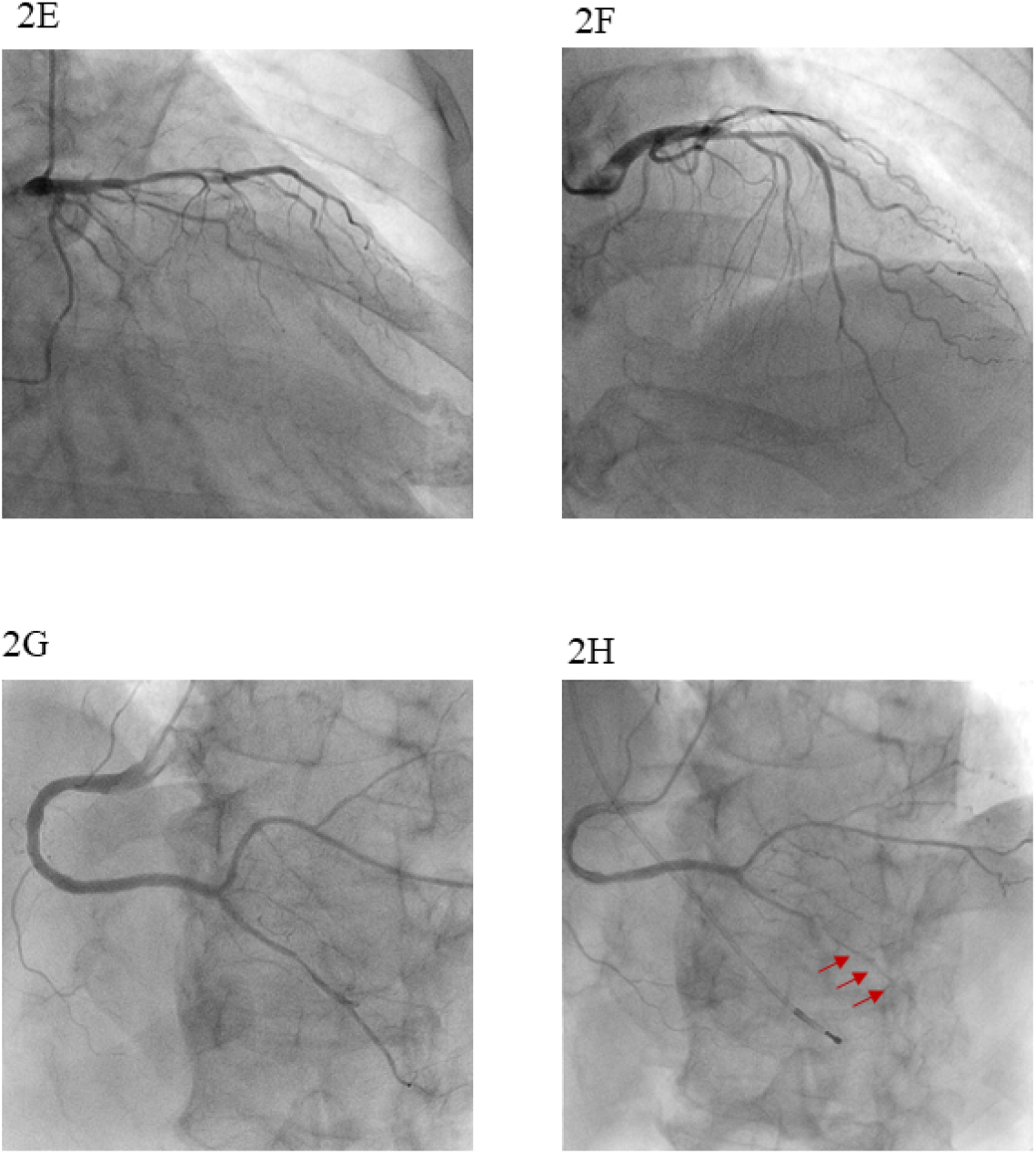
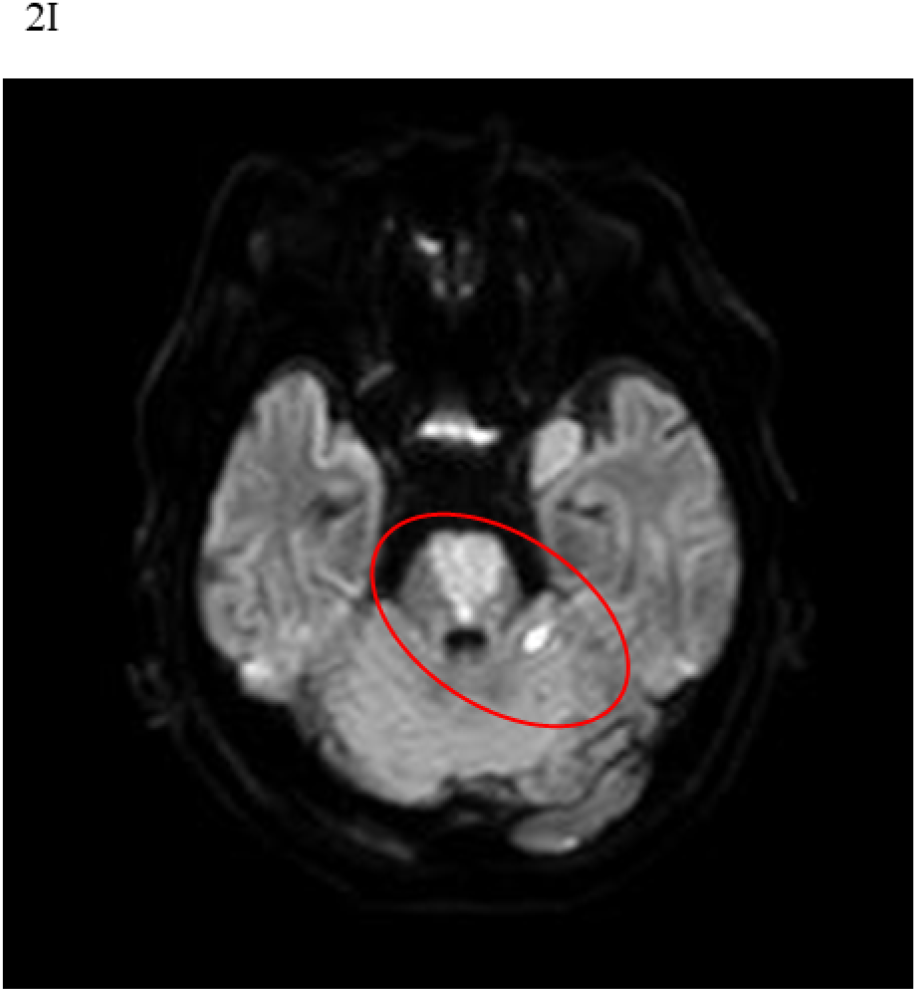
Representative case. A 76-year-old male suffered from squeezing chest pain during sleep. Flow-mediated dilation was moderately reduced (5.80%). Coronary computed tomography angiography showed a moderate degree of atherosclerosis in major coronary arteries, including high-risk plaques (HRPs), despite no significant stenoses (26.8% of per cent atheroma volume [PAV] and HRPs in the proximal left descending artery) (2A–D). Invasive coronary angiography showed normal left coronary artery (2E and F) and right coronary artery (RCA) (2G), but coronary vasospasm was induced by 50 µg of acetylcholine in RCA (2H). An ischaemic stroke occurred 17 months later (2I). LMT-LAD, left main trunk to the left anterior descending artery; LCX, left circumflex artery

### Statistical Analysis

The Shapiro–Wilk test was used to assess the normality of continuous data. Variables with a normal distribution are expressed as mean values ± standard deviation, and asymmetrically distributed data are presented as medians and interquartile ranges. Categorical variables are presented as frequencies (percentages). Differences between the two groups were evaluated using the Mann–Whitney U test or Student’s t-test for continuous variables and the chi-squared test for categorical variables. Kaplan-Meier survival analysis was used to estimate the interval from CTA to MACCE. Differences in the time-to-event curves were compared using log-rank statistics.

The effect of variables on MACCE was evaluated using univariate Cox proportional hazards models. Age, sex, and other variables (P<0.05) in the univariate analysis were included in the multivariate Cox proportional hazard analysis. Results are reported as hazard ratios (HRs) with 95% confidence intervals (CIs). The increased discriminative value after adding PAV, HRPs, and FMD to the baseline model, including age, sex, and hypertension, was estimated using the C-index, net reclassification improvement (NRI), and integrated discrimination improvement (IDI).

The C-index was defined as the area under the receiver operating characteristic (ROC) curve (AUC) between individual predicted probabilities and the confirmed occurrence of MACCE^19^. NRI indicated how many patients improved their predicted probabilities for MACCE occurrence. IDI represented the average improvement in predicted probabilities for MACCE after adding new data into the baseline model^20^. The interclass correlation coefficient was used to determine the intra- and inter-observer reproducibility for the measurement of PAV and the presence of HRP in 20 randomly selected patients. The analysis was conducted using JMP version 13 (SAS institute, Cary, NC, USA), and P-values <0.05 were considered statistically significant.

## Results

### Patient Characteristics

The mean FMD in 154 patients (mean age: 61.0±12.9 years, 89 males) was 6.3±1.7%, and HRPs were detected in 18 patients (11.7%). The median and mean CACS were 0 (IQR: 0– 82.8) and 105.7±250.1, respectively, and the mean PAV was 22.0±4.6% (Table 1). FMD tended to be lower in patients with HRPs than in those without HRPs (5.6±1.6% vs 6.4±1.7%, P=0.057). FMD was also moderately correlated with PAV (r=0.275, P<0.01; Figure 3). Blinded assessment for HRPs and PAV in each vessel was performed in a randomly selected, representative subgroup of 20 patients. The kappa statistics for intra- and inter-observer variabilities were 0.95 and 0.90 for HRPs, and 0.88 and 0.82 for PAV, respectively.

**Figure 3.**
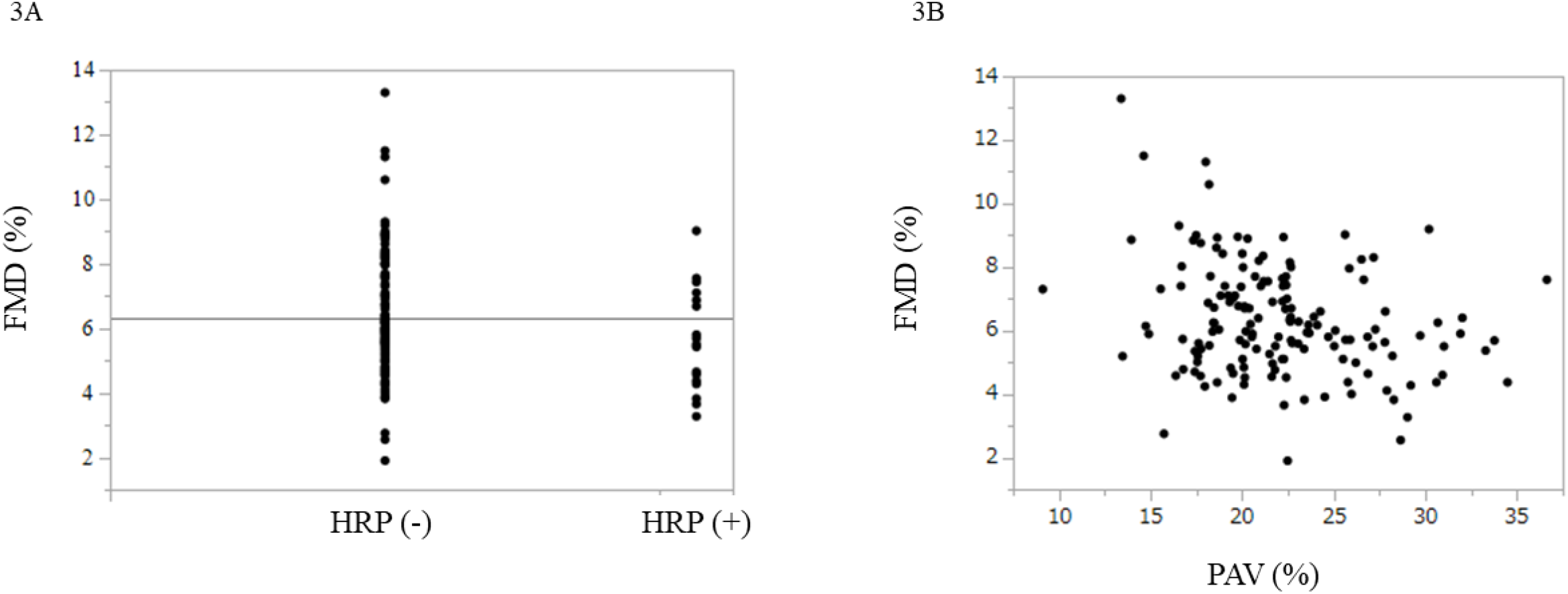
Relationship between HRPs, PAV, and FMD. FMD, flow-mediated dilation; PAV, per cent atheroma volume; HRP, high-risk plaque

**Table 1.**
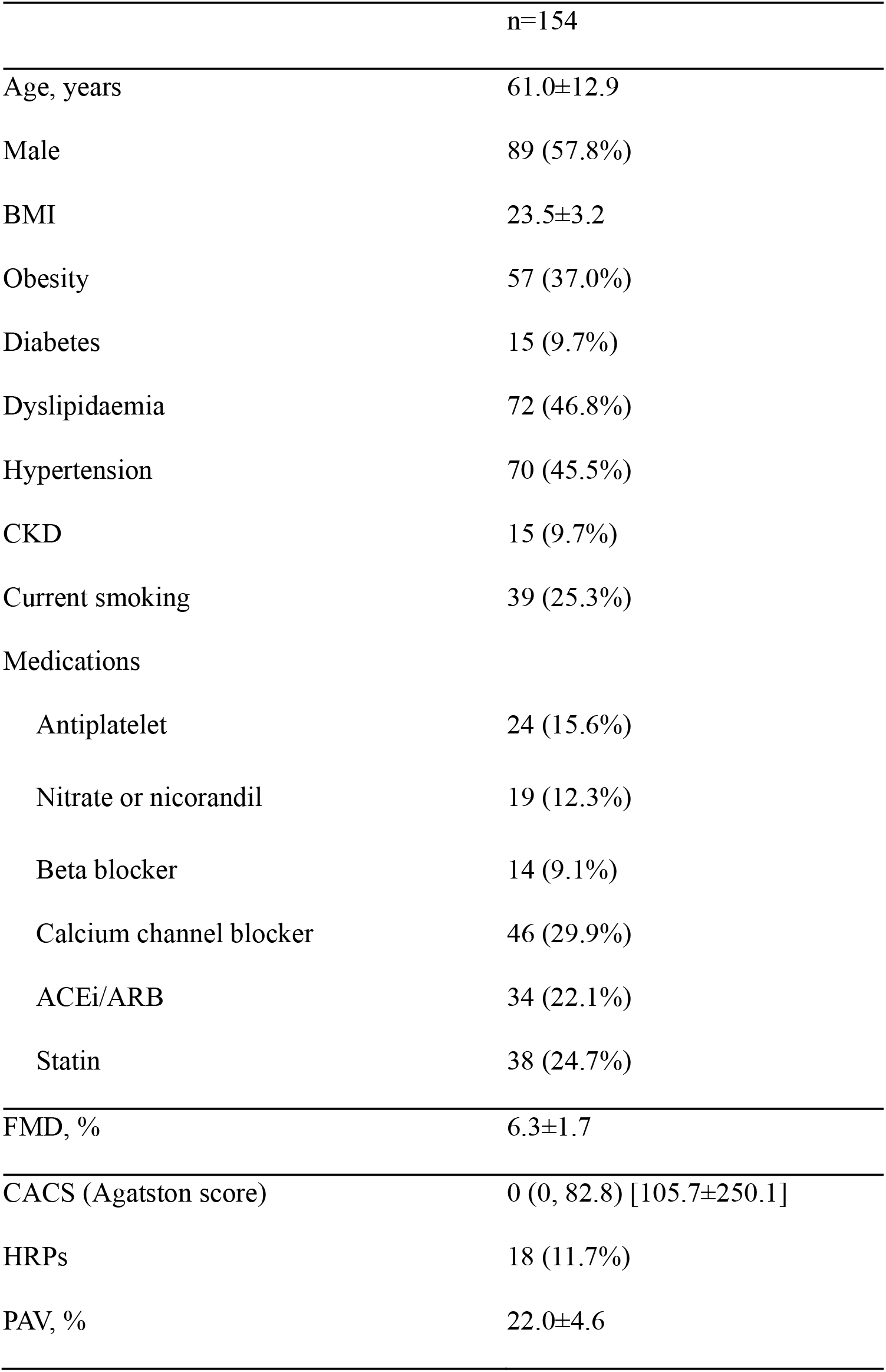
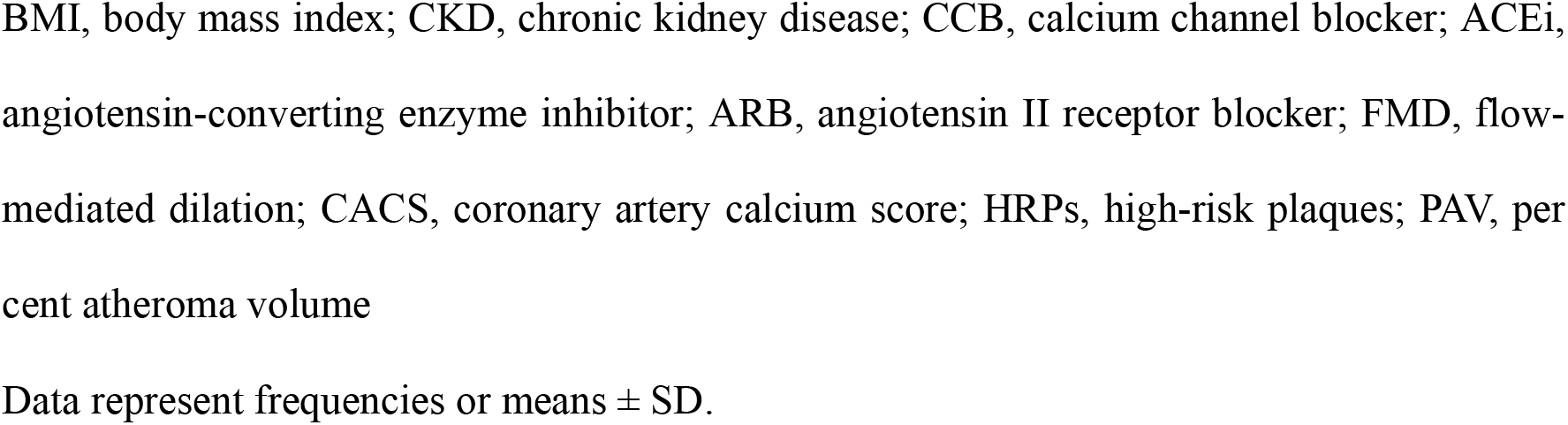
Patient background

### Cumulative Incidence of MACCE

The mean follow-up period was 4.7±2.7 years. Within the follow-up period, MACCE occurred in 19 patients: five with ACS, one resuscitation from ventricle fibrillation, seven with late revascularisation due to the progression of angina pectoris, and six with cerebral infarctions. Univariate analysis of MACCE occurrence revealed significant differences in hypertension, FMD, PAV, and the presence of HRPs (Table 2). After adjusting for age, sex, and hypertension, FMD, PAV, and HRPs were found to be independent predictors of MACCE.

**Table 2.**
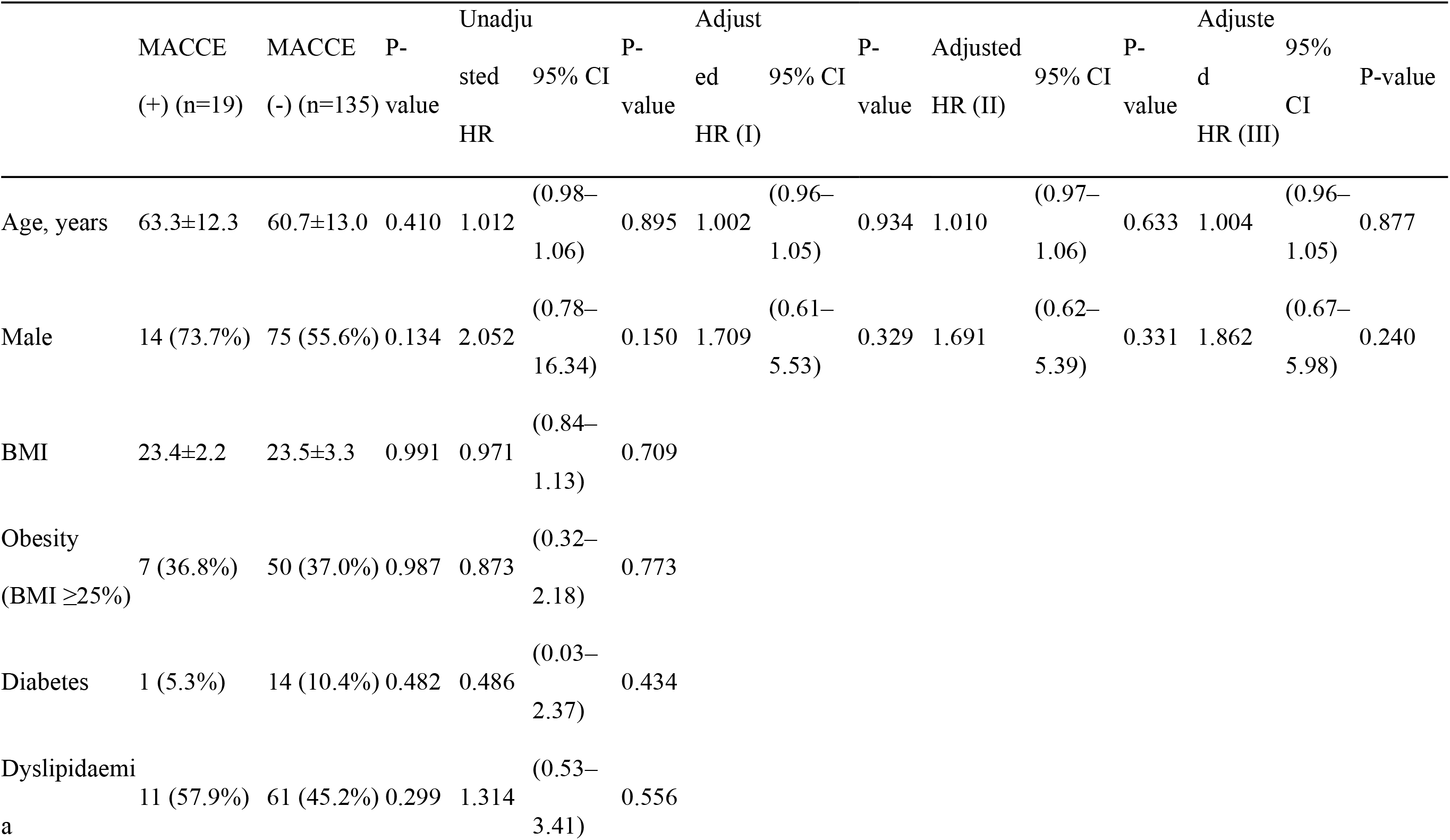
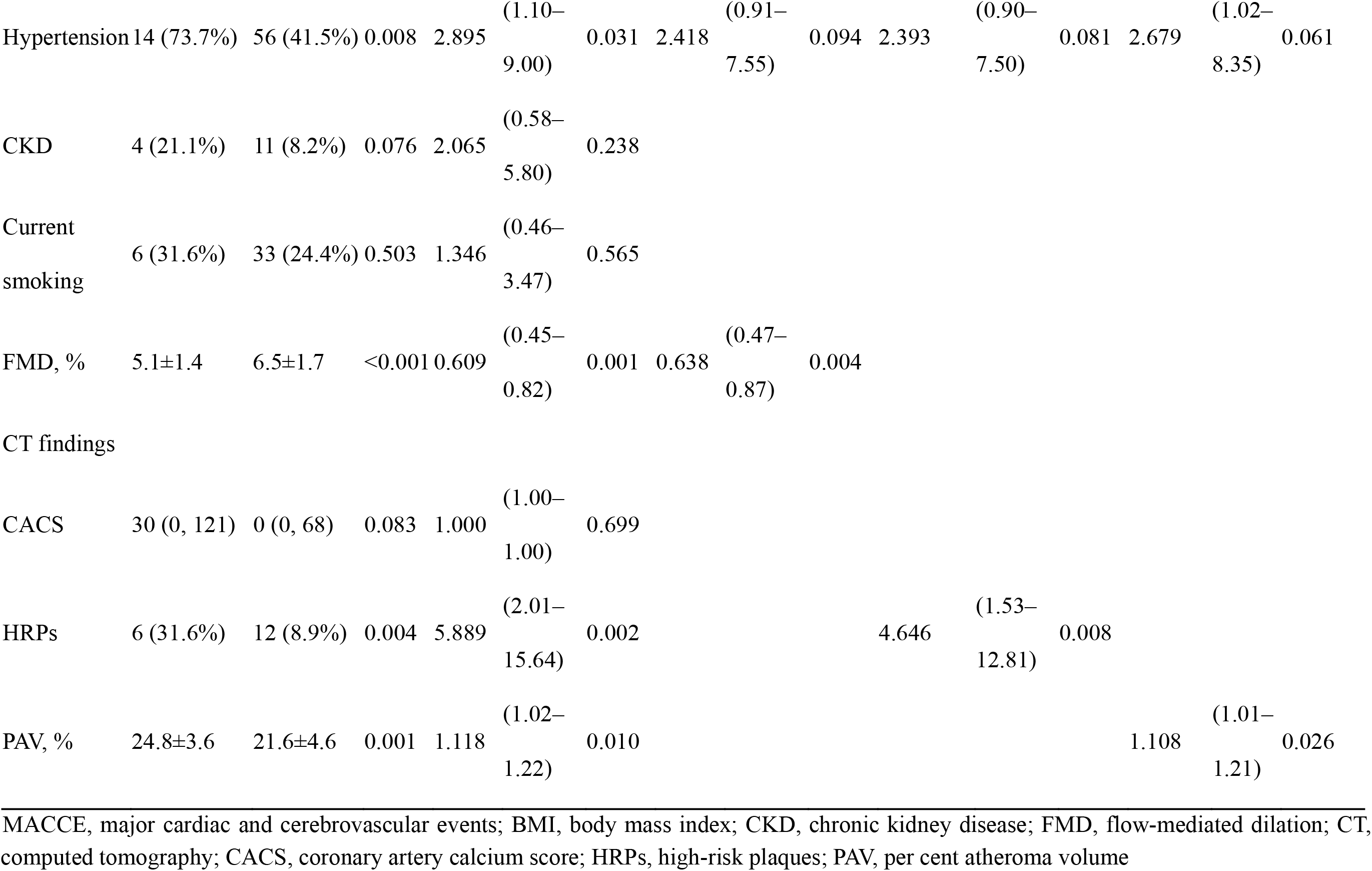
Univariate and multivariate analysis for MACCE

MACCE occurred more in patients with FMD <6.0% than in those with FMD ≥6.0% (Log-rank P=0.009). Similarly, the MACCE occurrence rate was higher in those with PAV ≥21.0% than in those with PAV <21.0% (Log-rank P=0.016). The MACCE occurrence rate was also higher in those with HRPs than in those without HRPs (Log-rank P<0.001; Figure 4 A–C). Patients were stratified into four groups based on the following factors: FMD <6.0%, PAV ≥21.0%, and the presence of HRPs. Cumulative incidence curves are shown in Figure 5. Compared with the patients with 0 points, hazard ratios of those with 1, 2, and 3 points were 2.76 (95% CI: 0.41–54.03, P=0.322), 9.89 (95% CI: 1.89–181.44, P=0.004), and 28.43 (95% CI: 4.15–559.55, P<0.001), respectively.

**Figure 4.**
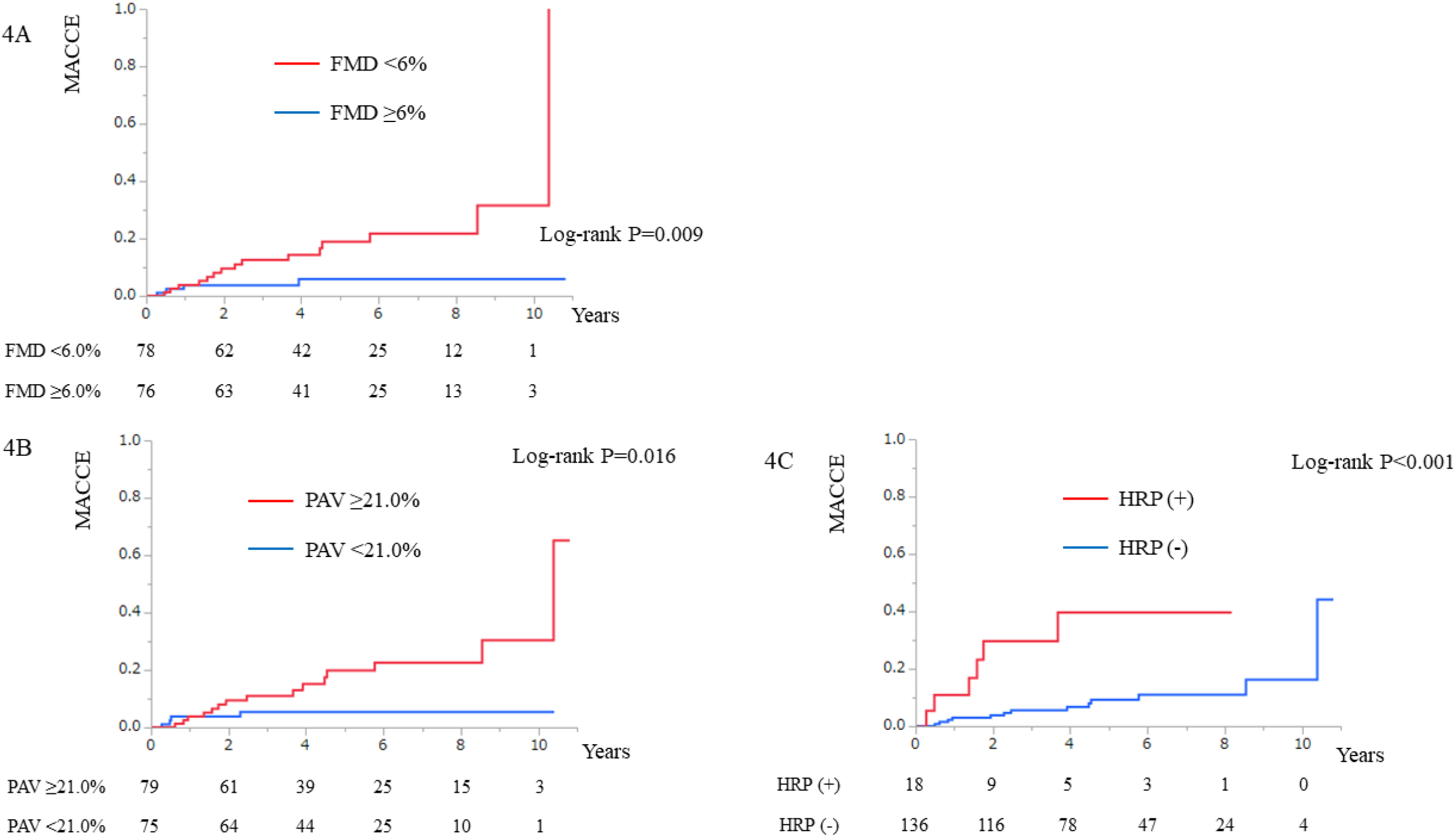
Kaplan–Meier curves for MACCE on FMD, PAV, and the presence of HRPs. MACCE, major cardiac and cerebrovascular events; FMD, flow-mediated dilation; PAV, per cent atheroma volume; HRP, high-risk plaque

**Figure 5.**
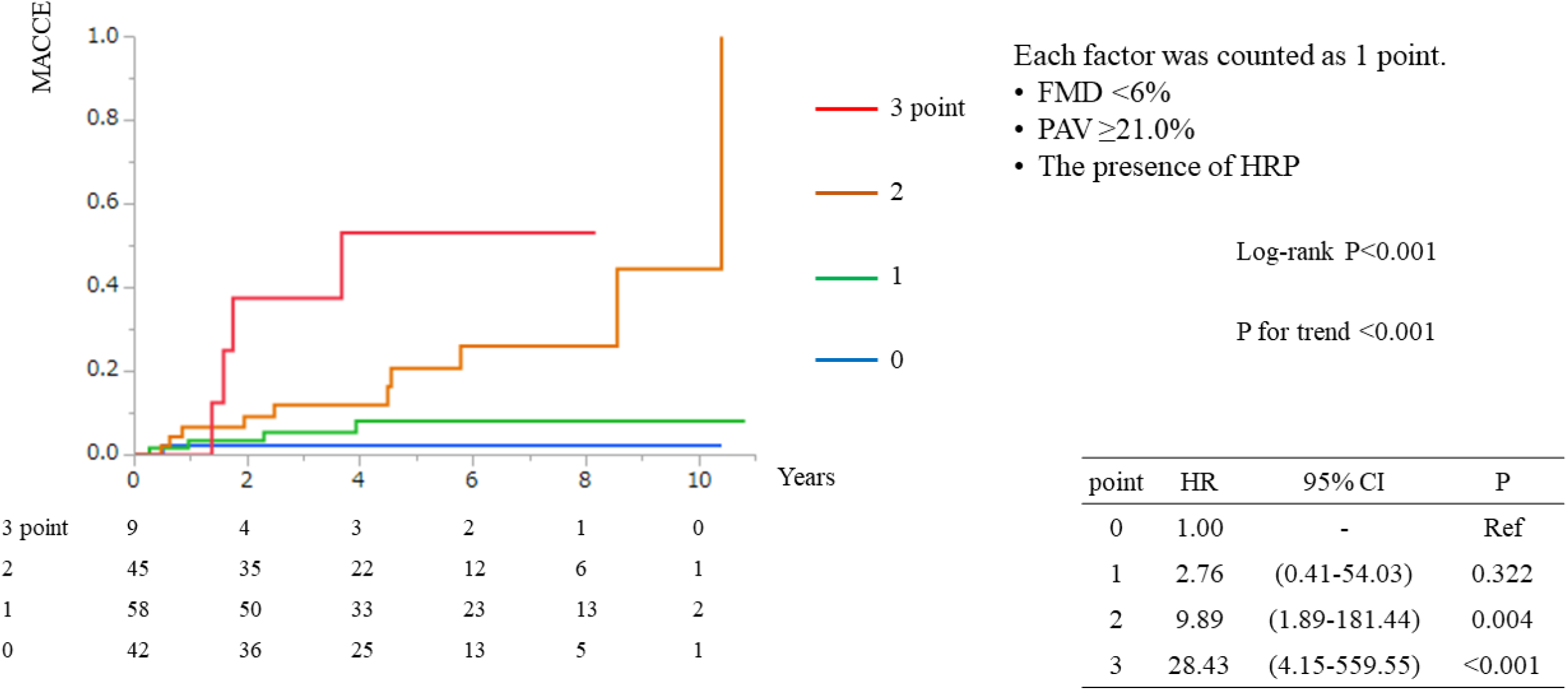
Kaplan–Meier curves for MACCE on the combination of FMD and CT findings. MACCE, major cardiac and cerebrovascular events; FMD, flow-mediated dilation; PAV, per cent atheroma volume; HRP, high-risk plaque

### Model Discrimination

The incremental prognostic value of FMD, PAV, the presence of HRPs, and their combination was tested against a baseline model including age, sex, and hypertension. Adding FMD, PAV, and HRPs to the baseline model improved the C-index (0.712–0.831, P=0.023). Adding the CT findings (PAV and HRPs) to the model including the baseline and FMD, improved the NRI (0.462, P=0.030) and IDI (0.046, P=0.041; Table 3).

**Table 3.** Model discrimination for MACCE

|  | C-index<br>(95% CI) | P-value | NRI | P-value | IDI | P-value |
| --- | --- | --- | --- | --- | --- | --- |
| (A) Baseline | 0.712<br>(0.571–0.854) | - | Ref | - | Ref | - |
| (B) Baseline<br>+ FMD | 0.783<br>(0.663–0.904) | 0.0672 | 0.537 | 0.014 | 0.066 | 0.009 |
| (C) Baseline<br>+ PAV | 0.774<br>(0.656–0.892) | 0.1028 | 0.626 | 0.005 | 0.036 | 0.012 |
| (D) Baseline<br>+ HRPs | 0.753<br>(0.616–0.891) | 0.2937 | 0.531 | 0.015 | 0.033 | 0.093 |
| (E) Baseline<br>+ FMD + PAV + HRPs | 0.831<br>(0.732–0.930) | 0.0230 | 1.032 | <0.001 | 0.111 | <0.001 |
| (E) vs (B) | - | 0.1332 | 0.462 | 0.030 | 0.046 | 0.041 |
| (E) vs (C) | - | 0.2064 | 0.777 | 0.001 | 0.076 | 0.010 |
| (E) vs (D) | - | 0.0669 | 0.973 | <0.001 | 0.079 | 0.006 |

MACCE, major cardiac and cerebrovascular events; NRI, net reclassification improvement; IDI, integrated discrimination improvement; FMD, flow-mediated dilation; PAV, per cent atheroma volume; HRP, high-risk plaque

## Discussion

The present study demonstrated that brachial FMD was weakly correlated with PAV and the presence of HRPs in the coronary artery tree. Both FMD and CT findings were useful for predicting cardiovascular events in patients, and the combination of these findings improved their prognostic value. FMD is an indicator of endothelial function and preclinical atherosclerosis. The quantitative coronary plaque volume on CTA estimates the extent of coronary atherosclerosis, and the presence of HRPs indicates the patient’s vulnerability to cardiovascular events. To the best of our knowledge, this study is the first to show that combining premature coronary atherosclerosis and endothelial function aids the stratification of patients’ cardiovascular risk.

Several studies have demonstrated the prognostic value of brachial FMD for cardiovascular events^2-4^. However, FMD has failed to demonstrate an added value to classical cardiovascular risk factors in terms of model discrimination^21, 22^. Furthermore, FMD may only reflect the compound risk burden that impacts vessel function and not provide incremental risk prediction^2^.

We have previously reported that persons with HRPs are at risk of future coronary events, including ACS or late revascularisation, due to the progression of coronary artery stenosis^9-11^. Meanwhile, automated software-based quantification of the total coronary plaque volume has emerged^6-8^ and is considered one of the most accurate methods for assessing the coronary artery disease (CAD) burden. Deseive et al. showed the long-term predictive value of the quantified total plaque volume using CTA^8^.

Arakawa et al. examined FMD and CACS in patients diagnosed with significant coronary artery stenoses and scheduled for invasive coronary angiography. Since the combined measurement of FMD and CACS identified the multi-vessel disease and high SYNTAX scores with high accuracy, they concluded that endothelial dysfunction and CACS might provide complementary information for predicting the extent and severity of CAD^16^. However, they studied only patients with significant coronary artery stenosis and did not perform quantitative analyses of the coronary artery tree. Lakshmanan et al. retrospectively examined 100 patients with intermediate cardiac risk and atypical symptoms who underwent FMD and coronary CTA. They concluded that FMD was independently associated with the presence and extent of subclinical atherosclerosis on CTA. However, they evaluated the extent of coronary atherosclerosis using segment scoring and did not quantify the total plaque volume. More than half of the patients had CACS >100. In addition, more patients with advanced atherosclerosis were registered in their study than in the present study^15^.

In the present study, FMD was weakly correlated with total plaque volume on CTA. In the population with subclinical coronary atherosclerosis, after excluding patients with significant coronary artery stenoses, the atherosclerotic plaque burden, presence of vulnerable plaque, and endothelial function did not correspond, although there were correlations. This study demonstrates the usefulness of these combinations for predicting cardiovascular events.

## Limitations

This study had several limitations. First, this was a retrospective, single-centre study with a small sample size. Patients who underwent both brachial FMD and coronary CTA for suspected CADs were enrolled. Thus, there is a possibility of patient selection bias. Furthermore, the FMD data and coronary CTA findings will likely affect patient management. Second, there was no precise assessment of plaque volume or characteristics using tools such as IVUS. However, many studies have validated the use of semi-quantitative and plaque assessments. This study obtained excellent intra- and inter-observer reproducibility for CTA plaque characterisation and quantification.

## Conclusions

The present study shows that brachial FMD, reflecting endothelial function, weakly correlates with the extent of plaque burden or HRPs in the coronary artery tree. Although FMD and CTA findings are useful in predicting cardiovascular events in patients, their combination synergises their prognostic abilities.

## Data Availability

All data generated or analyzed during this study are included in this published article and its supplementary information files. The datasets used and/or analyzed during the current study are available from the corresponding author upon reasonable request.

## Clinical Perspective

- This study is the first to show that combining plaque volume and characteristics observed on computed tomography (CT) angiography and flow-mediated dilation enables cardiovascular risk stratification in patients without a history of cardiovascular disease.
- Combining non-invasive endothelial function tests and CT findings of major coronary arteries is useful for stratifying cardiovascular risk among patients with subclinical coronary atherosclerosis.

## Non-standard Abbreviations and Acronyms

ACS: acute coronary syndrome
AUC: area under the ROC curve
CACS: coronary artery calcium score
CAD: coronary artery disease
CI: confidence interval
CT: computed tomography
CTA: computed tomography angiography
FMD: flow-mediated dilation
HR: hazard ratio
HRP: high-risk plaque
HU: Hounsfield unit
IDI: integrated discrimination improvement
IVUS: intravascular ultrasound
NRI: net reclassification improvement
MACCE: major cardiac and cerebrovascular events
PAV: per cent atheroma volume
ROC: receiver operating characteristic

## Acknowledgements

None

